# SARS-CoV-2 and Stroke Characteristics: A Report from the Multinational COVID-19 Stroke Study Group

**DOI:** 10.1101/2020.08.05.20169169

**Authors:** Shima Shahjouei, Georgios Tsivgoulis, Ghasem Farahmand, Eric Koza, Ashkan Mowla, Alireza Vafaei Sadr, Arash Kia, Alaleh Vaghefi Far, Stefania Mondello, Achille Cernigliaro, Annemarei Ranta, Martin Punter, Faezeh Khodadadi, Mirna Sabra, Mahtab Ramezani, Soheil Naderi, Oluwaseyi Olulana, Durgesh Chaudhary, Aicha Lyoubi, Bruce Campbell, Juan F. Arenillas, Daniel Bock, Joan Montaner, Saeideh Aghayari Sheikh Neshin, Diana Aguiar de Sousa, Matthew S. Tenser, Ana Aires, Merccedes De Lera Alfonso, Orkhan Alizada, Elsa Azevedo, Nitin Goyal, Zabihollah Babaeepour, Gelareh Banihashemi, Leo H. Bonati, Carlo Cereda, Jason J. Chang, Miljenko Crnjakovic, GianMarco De Marchis, Massimo Del Sette, Seyed Amir Ebrahimzadeh, Mehdi Farhoudi, Ilaria Gandoglia, Bruno Gonçalves, Christoph Griessenauer, Mehmet Murat Hancı, Aristeidis H. Katsanos, Christos Krogias, Ronen Leker, Lev Lotman, Jeffrey Mai, Shailesh Male, Konark Malhotra, Branko Malojcic, Teresa Mesquita, Asadollah Mirghasemi, Hany Mohamed Aref, Zeinab Mohseni Afshar, Jusun Moon, Mika Niemelä, Behnam Rezaei Jahromi, Lawrence Nolan, Abhi Pandhi, Jong-Ho Park, João Pedro Marto, Francisco Purroy, Sakineh Ranji-Burachaloo, Nuno Reis Carreira, Manuel Requena, Marta Rubiera, Seyed Aidin Sajedi, João Sargento-Freitas, Vijay Sharma, Thorsten Steiner, Kristi Tempro, Guillaume Turc, Yassaman Ahmadzadeh, Mostafa Almasi-Dooghaee, Farhad Assarzadegan, Arefeh Babazadeh, Humain Baharvahdat, Fabricio Cardoso, Apoorva Dev, Mohammad Ghorbani, Ava Hamidi, Zeynab Sadat Hasheminejad, Sahar Hojjat-Anasri Komachali, Fariborz Khorvash, Firas Kobeissy, Hamidreza Mirkarimi, Elahe Mohammadi-Vosough, Debdipto Misra, Ali Reza Noorian, Peyman Nowrouzi-Sohrabi, Sepideh Paybast, Leila Poorsaadat, Mehrdad Roozbeh, Behnam Sabayan, Saeideh Salehizadeh, Alia Saberi, Mercedeh Sepehrnia, Fahimeh Vahabizad, Thomas Yasuda, Ahmadreza Hojati Marvast, Mojdeh Ghabaee, Nasrin Rahimian, Mohammad Hossein Harirchian, Afshin Borhani-Haghighi, Rohan Arora, Saeed Ansari, Venkatesh Avula, Jiang Li, Vida Abedi, Ramin Zand

## Abstract

**Background:** Stroke is reported as a consequence of SARS-CoV-2 infection. However, there is a lack of regarding comprehensive stroke phenotype and characteristics

**Methods:** We conducted a multinational observational study on features of consecutive acute ischemic stroke (AIS), intracranial hemorrhage (ICH), and cerebral venous or sinus thrombosis (CVST) among SARS-CoV-2 infected patients. We further investigated the association of demographics, clinical data, geographical regions, and countries’ health expenditure among AIS patients with the risk of large vessel occlusion (LVO), stroke severity as measured by National Institute of Health stroke scale (NIHSS), and stroke subtype as measured by the TOAST criteria. Additionally, we applied unsupervised machine learning algorithms to uncover possible similarities among stroke patients.

**Results:** Among the 136 tertiary centers of 32 countries who participated in this study, 71 centers from 17 countries had at least one eligible stroke patient. Out of 432 patients included, 323(74.8%) had AIS, 91(21.1%) ICH, and 18(4.2%) CVST. Among 23 patients with subarachnoid hemorrhage, 16(69.5%) had no evidence of aneurysm. A total of 183(42.4%) patients were women, 104(24.1%) patients were younger than 55 years, and 105(24.4%) patients had no identifiable vascular risk factors. Among 380 patients who had known interval onset of the SARS-CoV-2 and stroke, 144(37.8%) presented to the hospital with chief complaints of stroke-related symptoms, with asymptomatic or undiagnosed SARS-CoV-2 infection. Among AIS patients 44.5% had LVO; 10% had small artery occlusion according to the TOAST criteria. We observed a lower median NIHSS (8[3-17], versus 11 [5-17]; p=0.02) and higher rate of mechanical thrombectomy (12.4% versus 2%; p<0.001) in countries with middle to high-health expenditure when compared to countries with lower health expenditure. The unsupervised machine learning identified 4 subgroups, with a relatively large group with no or limited comorbidities.

**Conclusions:** We observed a relatively high number of young, and asymptomatic SARS-CoV-2 infections among stroke patients. Traditional vascular risk factors were absent among a relatively large cohort of patients. Among hospitalized patients, the stroke severity was lower and rate of mechanical thrombectomy was higher among countries with middle to high-health expenditure.

## Introduction

Since the emergence of the Coronavirus disease 2019 (COVID-19) pandemic, several cases of cerebrovascular events were reported among patients with SARS-CoV-2.[1,2] Studies presented the incidence and prevalence of acute ischemic stroke (AIS), intracranial hemorrhage (ICH), and cerebral venous or sinus thrombosis (CVST) in SARS-CoV-2 infected patients.[3-11] Majority of these reports focus on AIS patients. Some reports highlighted AIS in critically ill and older patients with a higher number of comorbidities,[1,6,12] some suggested a higher risk in younger and healthy individuals with male predilection,[3,7,13] and other reports compared these patients with the cohort of stroke patients who were not infected with the virus.[14,15]

Many studies proposed coagulopathy as the underlying pathophysiological mechanism for the cerebrovascular events.[15,16] Accordingly, studies suggested that stroke patients with SARS-CoV-2 present multiple cerebral infarcts,[8,12,13] systemic coagulopathies in multiple organs,[17] uncommon thrombotic events such as aortic[18] or common carotid artery thrombosis,[19] and simultaneous arterial and venous thrombus formation.[20] Small case series demonstrated a higher proportion of large vessel occlusions, [3,6,13], or cryptogenic strokes, [5,21] with elevated D-dimer level, liver enzymes, and inflammatory or renal failure biomarkers among the patients who experienced SARS-CoV-2 infection.[5,7,12,22] Besides, most of the studies noted a higher severity and mortality rate among stroke patients diagnosed with SARS-CoV-2 compared with others.[2,5,6,23,24]

We devised a multinational multiple phase study to evaluate the stroke risk[25] and characteristics among the SARS-CoV-2 infected patients. We investigated the association among demographics, clinical data, geographical regions, and countries health expenditure among AIS patients with the risk of large vessel occlusion (LVO), stroke severity as measured by National Institute of Health stroke scale (NIHSS),[26] and stroke subtype (documented using TOAST criteria – the Trial of Org 10172 in Acute Stroke Treatment).[27] We further applied unsupervised machine learning algorithms to uncover the possible similarities among these patients.

## Methods

### Study Design

The details of the study design are available in Supplemental Document 1. This multicenter, multinational observational study was conducted and reported according to the Strengthening the Reporting of Observational Studies in Epidemiology (STROBE),[28] and Enhancing the QUAlity and Transparency Of health Research (EQUATOR) guidelines. [29] The study protocol was designed by the investigators at the Neuroscience Institute of Geisinger Health System, Pennsylvania, USA, and received approval by the Institutional Review Board of Geisinger Health System and other participating institutions. Investigators from North America (Canada and six states of the United States), South America (Brazil and Mexico), Europe (Belgium, Croatia, Czech Republic, Finland, France, Germany, Greece, Ireland, Italy, Norway, Portugal, Spain, Sweden, and Switzerland), Asia and the Middle East (India, Iran, Iraq, Israel, Lebanon, Singapore, South Korea, Turkey, and the United Arab Emirates), Oceania (Australia and New Zealand), and Africa (Egypt, Nigeria, and Uganda) responded to our invitation. The centers were included by non-probability sampling and data were recruited until June 10th, 2020.

#### Participants

We included consecutive SARS-CoV-2 infected adult patients who had imaging confirmed subsequent stroke[30]— AIS, intracerebral hemorrhage, subarachnoid hemorrhage (SAH), and CVST. The preferred diagnostic criteria for SARS-CoV-2 was defined according to the World Health Organization (WHO) interim guidance.[31] Ischemic or hemorrhagic strokes were defined in the presence of a rapid onset of a neurological deficit with evidence of acute ischemic or hemorrhagic lesions on Computed Tomography (CT) or Magnetic Resonance Imaging (MRI). Patients who had transient stroke-like symptoms (transient ischemic attack, TIA) without acute lesions on CT or MRI were not included in this study due to the high diagnostic error. [32-34]

Patients who initially presented to the hospital with stroke-related chief complaints and asymptomatic SARS-CoV-2 infection, those who had a stroke while being hospitalized for SARS-CoV-2 infection, or patients with stroke-related admission who had confirmed prior diagnosis of SARS-CoV-2 were included in this study. We excluded patients with stroke incidents prior to the SARS-CoV-2 infection onset. The onset of SARS-CoV-2 was considered as either the symptoms onset or positive test, whichever was first.

#### Data Element and Processing

Collaborators were asked to provide data according to a core protocol. The age, sex, vascular risk factors and comorbidities (i.e., hypertension, diabetes mellitus, ischemic heart disease, atrial fibrillation, carotid stenosis, chronic kidney disease, cardiac ejection fraction <40%, active neoplasms, rheumatological diseases, smoking status, and history of TIA or stroke), and laboratory findings (i.e., the count for white blood cells, neutrophils, lymphocytes, and platelets, C-Reactive Protein, blood urea nitrogen, creatinine, alanine transaminase, aspartate transaminase, lactic acid dehydrogenase, fibrinogen, and D-dimer) were requested for the stroke patients. We also obtained additional data including the onset of the stroke and SARS-CoV-2 infection, the initiation of mechanical ventilation (if applicable), length of hospital stays, and patient disposition—still in the hospital, in-hospital death, being discharged to home, acute rehabilitation service, or nursing home. The details of neurological symptoms and investigations, imaging-based localization of the lesion(s), use of antiplatelets or anticoagulants prior to the stroke, and the National Institute of Health stroke scale (NIHSS) and the intracranial hemorrhage (ICH) score were also requested. In patients with AIS, beside TOAST criteria (the Trial of Org 10172 in Acute Stroke Treatment; defined as large artery atherosclerosis, cardioembolism, small artery occlusion, other determined etiology, and undetermined etiology)[27], the lesion(s) on diffusion-weighted imaging (DWI) or CT images were categorized as lacunar,[35] embolic/large vessel athero-thromboembolism, [36,37] vasculitis pattern, [38] or other phenotypes (borderzone or equivocal lesions). In this study, the AIS due to large-vessel occlusions (LVOs) are referred as occlusion of the internal carotid artery (ICA), middle cerebral artery (MCA) at M1 and M2, anterior cerebral artery (ACA) at A1, posterior cerebral artery (PCA) at P1, intracranial vertebral artery (VA), or basilar artery (BA).[39] Brain imaging findings were evaluated by local radiologists with expertise in neuroimaging.

We grouped the patients according to the age (younger versus older than 55-year-old, and younger versus older than 65-year-old),[40] comorbidities, laboratory findings, geographical regions (Middle East, Asia, Europe, and America), and countries’ health expenditure (low versus middle and high income based on WHO reports).[41] We considered the countries’ annual health expenditure of above US$1,000 (2015 to 2017, Supplemental Table 1) and total health expenditure of above US$10,000 (2010 to 2017, Supplemental Figure 1) as the cut-off. For the interval between SARS-CoV-2 infection and stroke, we grouped the patients as visiting the hospital with the chief complaint of strokelike symptoms and obscure viral infection versus those who presented the symptoms or were diagnosed by SARS-CoV-2 infection prior to the stroke.

#### Outcome measures

The outcome measures in this study were the presence versus absence of LVO, stroke severity as measured by NIHSS, and stroke subtype as measured by TOAST criteria. The severity of the stroke, according to NIHSS, was defined as no stroke symptoms (NIHSS=0), minor (NIHSS of 1 to 4), moderate (NIHSS of 5 to 15), moderate to severe (NIHSS of 16 to 20), and severe stroke (NIHSS 21-42).[26] We studied the outcome measures only among the patients with AIS. We did not analyze the disposition and length of stay as outcome measures since many patients were still in the acute phase or admitted in long-term acute care hospitals at the closure of our study.

#### Clustering and Subgroup Analyses

We performed clustering and subgroup analyses (Figure 1) to uncover the possible similarities among the patients in the entire cohort. Patients with AIS and intraparenchymal hemorrhage (IPH, hemorrhage exclusively within brain parenchyma) were grouped independently based on the reported comorbidities and laboratory findings with the aid of unsupervised machine learning algorithms (ML) or expert opinion (EX) (details available in Supplemental Document 2). In ML models, we used hierarchical and K-means clustering (ML-K) to group the patients into 2 to 5 subgroups (models ML-K_2_ to ML-K_5_). We also used the spectral clustering (ML-S) and clustered the patients into 2 to 5 subgroups (ML-S_2_ to ML-S_5_). The subgroup analysis considering the expert opinion (EX) defined the risk score as the sum of either all the 11 collected comorbidities (Expert-All, EX-A) or the 8 selected comorbidities (Expert-Selected, EX-S). We first divided the EX-A and EX-S into two subgroups (EX-A_2_ and EX-S_2_); subgroups “a” included the patients with zero or one comorbidity, and subgroup “b” included the patients with >1 comorbidity. We repeated the subgroup analysis by dividing the EX-A and EX-S into three subgroups (EX-A_3_ and EX-S_3_); we considered subgroup “a” as patients without any known comorbidity, subgroup “b” with one or two comorbidities, and subgroup “c” as the patients with >2 comorbidities. The above processes were repeated for the laboratory findings (data not shown); however, no patterns or significant differences among the subgroups were observed. The clustering was not performed for patients with SAH and CVST due to limited sample size.

**Figure 1.**
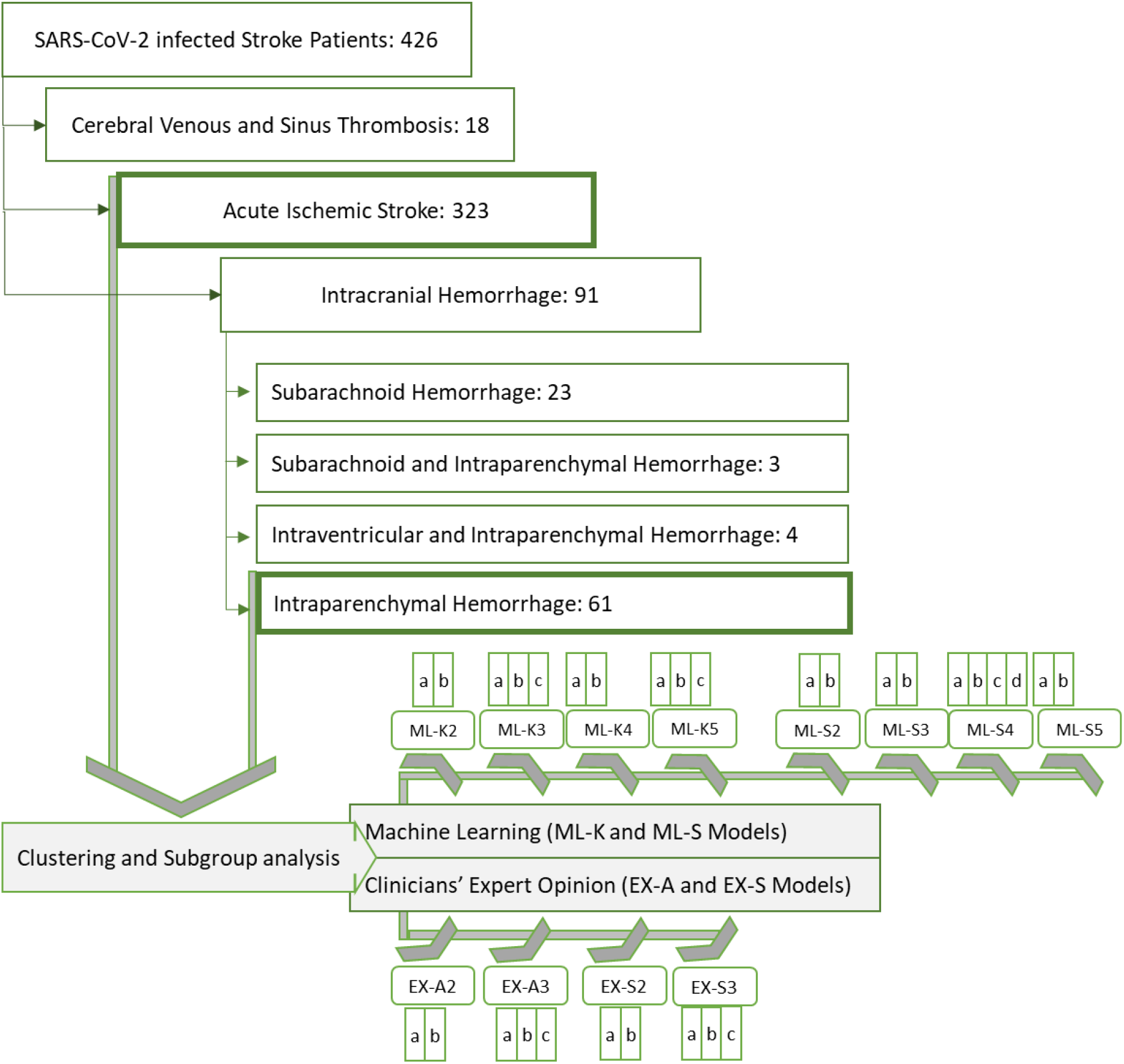
Flow chart. Overview of the included patients, approaches, and modeling.

#### Statistical Analysis and Modeling

We used descriptive statistics to summarize the data. Demographic data, comorbidities, laboratory findings, and neurological investigations were reported as medians and interquartile range (IQR), mean and standard deviations (SD), and under stratified categories when possible. The equality of the variances was assessed by Leven’s test. Categorical variables were reported as absolute frequencies and percentages. The comparisons between categorical variables were conducted with the Pearson chi-square test, while the differences among continuous variables were assessed by independent t-test and analysis of variance (ANOVA). A post-hoc z-test on the adjusted residuals, and Cramér’s phi, Tukey, or Dunnett’s tests were used to demonstrate the degree and direction of the associations in comparison of means, while post-hoc comparison of medians was conducted by Dunn-Bonferroni approach to compare subgroups. All tests were performed using IBM SPSS Statistics version 26[42], and p<0.05 was considered statistically significant. Bonferroni correction was used for adjusting all p values in multiple comparisons.

We used unsupervised ML algorithms to cluster the patients based on the comorbidities and laboratory findings. The laboratory values were scaled to 1-100 range and underwent Log_10_ transformation prior to the clustering. We applied hierarchical (complete linkage method) and k-means (Hartigan-Wong algorithm) clustering, and spectral clustering,[43] to produce 2 to 5 clusters. We used the contingency matrix (a.k.a contingency table)[44] to present the clusters of each model versus other models. The similarity of the models was calculated as follows:

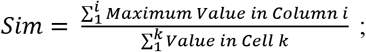 where *i* is the number of columns and k is the total number of cells in the contingency matrix. Similarities among the models were considered to be mild (50%-65%), moderate (65%-80%), and strong (80%-100%). Packages *stat[45]* and *gplots[46]* in R version 3.6.3, and scikit-learn package[47] in Python version 3.7, were used for machine learning algorithms and visualization.

### Results

Collaborators from 136 tertiary centers of 32 countries participated in this study. Among them, 71 centers from 17 countries had at least one stroke patient eligible for this study. One center in the Middle East could not provide data by the deadline. The rest of the centers did not have stroke patients who met our inclusion criteria (Supplemental Document 1). We received data on 432 patients—America:114(26.4%), Europe: 82(19.0%), Middle East: 228(52.8%), and Asia: 8(1.9%). Among them, 203(47.0%) patients were from countries with middle-to high-health expenditure. Overall, 323(74.8%) patients had AIS, 91(21.1%) ICH, and 18(4.2%) CVST.

The mean age for the entire cohort was 65.7±15.7 years. Out of 432 patients, majority were men--249(57.6%), p<0.001. Among 380 patients who had known interval onset of the SARS-CoV-2 and stroke, 144(37.8%) presented to the hospital with chief complaints of stroke-related symptoms, with asymptomatic or undiagnosed SARS-CoV-2 infection. In total, 105(24.4%) out of the 430 patients with complete comorbidity profiles had no identifiable vascular risk factor at the time of stroke incidence. Details of patient phenotype and demographic characteristics under each stroke subtype are presented in Table 1.

**Table 1.** The baseline characteristics, comorbidities, and laboratory findings among SARS-CoV-2 infected stroke.

| Parameter |  | Acute Ischemic Stroke<br>N = 323 (74.8%) | Intracerebral Hemorrhage<br>N = 68 (15.7%) | Subarachnoid<br>Hemorrhage<br>N = 23 (5.3%) | Cerebral<br>Venous/Sinus Thrombosis<br>N = 18 (4.2%) |
| --- | --- | --- | --- | --- | --- |
| Age; Mean (SD); Years |  | 67.2 ± 15.2 | 63 ± 16 | 62.6 ± 14.1 | 48.9 ± 11.5 |
| Age; Median [IQR]; Years |  | 68 [58 – 78] | 65 [54 – 75] | 62 [54 – 74] | 51 [39.5 – 55.2] |
|  | <40; N (%) | 23 (7.1) | 8 (12.9) | 1 (4.3) | 5 (27.8) |
|  | 41-64; N (%) | 110 (34.1) | 20 (32.3) | 12 (52.2) | 11 (61.1) |
|  | 65-74; N (%) | 77 (23.8) | 13 (21) | 5 (21.7) | 2(11.1) |
|  | ≥75; N (%) | 113 (53) | 21 (33.9) | 5 (21.7) | 0 (0) |
| Sex; Women; N (%) |  | 130 (40.2) | 30 (44.1) | 12 (52.2) | 11 (61.1) |
| Stroke Symptoms as the Chief Complaint; N (%) |  | 104 (36.1) | 31 (53.4) | 8 (36.4) | 4 (22.2) |
| SARS-CoV-2 to Stroke Interval; Mean (SD); Days |  | 5 ± 7 | 5 ± 8 | 3 ± 5 | 3 ± 4 |
| SARS-CoV-2 to Stroke Interval; Median [IQR]; Days |  | 3 [0-9] | 0 [0-10.3] | 1.5 [0-5.3] | 4.5 [0.7-10.3] |
| National Institutes of Health Stroke Scale (NIHSS); Median [IQR] |  | 9 [4-17] | 10 [0-19] | 6 [0-16] | 14 [7-17] |
| Intracerebral Hemorrhage (ICH) Score; Median [IQR] |  | - | 3 [2 - 4] | - | - |
| Mechanical Ventilation; N (%) |  | 238 (73.7) | 31 (50) | 12 (52.2) | 10 (55.6) |
| Initiation of Mechanical Ventilation; Median [IQR]; Days |  | 2 [1-3] | 2 [1-4] | 5 [1-15] | 1 [1-2] |
| Length of Hospital Stay; Mean (SD); Days |  | 12 ± 12 | 15 ± 18 | 18 ± 20 | 18 ± 14 |
| Length of Hospital Stay; Median [IQR]; Days |  | 7 [4-16] | 8 [3-21] | 11 [4.2-31] | 14 [7-30] |
| Disposition* |  |  |  |  |  |
| Discharged Home; N (%) |  | 127 (42.8) | 10 (17.9) | 3 (15) | 12 (66.7) |
| In-Hospital Mortality; N (%) |  | 82 (27.6) | 35 (62.5) | 11 (55) | 3 (16.7) |
| Still in Hospital, or Dispositioned to Subacute Care; N (%) |  | 88 (29.6) | 11 (22) | 6 (30) | 3 (16.7) |
| Medications |  |  |  |  |  |
| Prior Anti-Platelet Therapy; N (%) |  | 87 (28.0) | 16 (28.1) | 3 (14.3) | 0 (0.0) |
| Prior Anti-Coagulant Therapy; N (%) |  | 28 (9.0) | 6 (10.5) | 2 (9.5) | 4 (23.5) |
| Region |  |  |  |  |  |
| Middle East; N (%) |  | 153 (47.4) | 43 (63.2) | 17 (73.9) | 15 (83.3) |
| America; N (%) |  | 6 (1.9) | 0 (0.0) | 0 (0.0) | 2 (11.1) |
| Europe; N (%) |  | 88 (27.2) | 19 (27.9) | 6 (26.1) | 1 (5.6) |
| Asia; N (%) |  | 76 (23.5) | 6 (8.8) | 0 (0.0) | 0 (0.0) |
| Countries' Health Expenditure |  |  |  |  |  |
| Middle to High; N (%) |  | 170 (52.6) | 26 (38.2) | 5 (21.7) | 2 (11.1) |
| Low; N (%) |  | 153 (47.4) | 42 (61.8) | 18 (78.3) | 16 (88.9) |
| Comorbidities |  |  |  |  |  |
| Hypertension; N (%) |  | 202 (63.1) | 43 (63.2) | 14 (60.9) | 1 (5.6) |
| Diabetes Mellitus; N (%) |  | 111 (34.6) | 21 (30.9) | 4 (17.4) | 2 (11.1) |
| Ischemic Heart Disease; N (%) |  | 72 (24.3) | 12 (17.6) | 4 (21.1) | 0 (0) |
| Atrial Fibrillation; N (%) |  | 45 (14.1) | 3 (4.4) | 2 (8.7) | 0 (0) |
| Carotid Stenosis; N (%) |  | 38 (12.8) | 2 (2.9) | 0 (0) | 0 (0) |
| Chronic Kidney Disease; N (%) |  | 42 (13.1) | 9 (13.2) | 3 (13) | 2 (11.1) |
| Cardiac Ejection Fraction <40%; N (%) |  | 24 (8.1) | 7 (10.3) | 0 (0) | 0 (0) |
| Active Neoplasm; N (%) |  | 21 (7.1) | 4 (5.9) | 1 (4.3) | 0 (0) |
| Rheumatological Disease; N (%) |  | 5 (1.7) | 0 (0) | 1 (4.3) | 0 (0) |
| Prior Stroke or Transient Ischemic Attack; N (%) |  | 5 (1.7) | 0 (0) | 0 (0) | 0 (0) |
| Smoking; N (%) |  | 53 (16.6) | 11 (16.2) | 3 (13) | 0 (0) |

| Parameter |  | Acute Ischemic Stroke<br>N = 323 (74.8%) | Intracerebral Hemorrhage<br>N = 68 (15.7%) | Subarachnoid Hemorrhage<br>N = 23 (5.3%) | Cerebral Venous/Sinus Thrombosis<br>N = 18 (4.2%) |
| --- | --- | --- | --- | --- | --- |
| Laboratory findings |  |  |  |  |  |
| White Blood Cell Count; Mean (SD); x109/L |  | 9.8 ± 4.8 | 11.4 ± 10.0 | 13.5 ± 12.9 | 11.1 ± 6.0 |
| White Blood Cell Count; Median [IQR]; x109/L |  | 9 [6.8 – 11.2] | 9.5 [7.7 – 12.1] | 10 [8.8 – 12.2] | 11.1 [6.7 – 14.8] |
|  | <4 x109/L; N (%) | 7 (2.3) | 3 (5.7) | 0 (0) | 2 (22.2) |
|  | 4-10 x109/L; N (%) | 184 (61.1) | 29 (54.7) | 10 (50) | 4 (44.4) |
|  | 10-20 x109/L; N (%) | 96 (31.9) | 18 (34) | 9 (45) | 3 (33.3) |
|  | ≥20 x109/L; N (%) | 14 (4.7) | 5 (10) | 1 (5) | 0 (0) |
| Neutrophil Count; Mean (SD); x109/L |  | 7.7 ± 4.5 | 9.3 ± 8.5 | 9.6 ± 6.1 | 9.2 ± 5.1 |
| Neutrophil Count; Median [IQR]; x109/L |  | 6.8 [4.8 – 9.2] | 7.2 [5.2 – 10.6] | 7.3 [5.4 – 11] | 9.4 [4.7 – 14.2] |
|  | <4 x109/L; N (%) | 43 (15.2) | 5 (10) | 1 (5) | 1 (12.5) |
|  | 4-10 x109/L; N (%) | 182 (64.3) | 33 (66) | 12 (60) | 6 (75) |
|  | 10-20 x109/L; N (%) | 49 (17.3) | 10 (20) | 6 (30) | 1 (12.5) |
|  | ≥20 x109/L; N (%) | 9 (3.2) | 2 (4) | 1 (5) | 0 (0) |
| Lymphocyte Count; Mean (SD); x109/L |  | 1.5 ± 1.5 | 2.0 ± 3.6 | 2.8 ± 6.3 | 2.0 ± 1.3 |
| Lymphocyte; Median [IQR]; x109/L |  | 1.3 [0.9 – 1.9] | 1.3 [0.9– 2.0] | 1.3 [0.9-1.9] | 1.9 [1.2 – 2.3] |
|  | <1 x109/L; N (%) | 93 (32.5) | 21 (39.6) | 6 (28.6) | 1 (12.5) |
|  | 1-2 x109/L; N (%) | 130 (45.5) | 19 (35.8) | 10 (47.6) | 4 (50) |
|  | 2-3 x109/L; N (%) | 41 (14.3) | 9 (35.8) | 3 (14.3) | 2 (25) |
|  | 3-4 x109/L; N (%) | 11 (3.8) | 1 (1.9) | 0 (0) | 1 (12.5) |
|  | ≥4 x109/L; N (%) | 11 (3.8) | 3 (5.7) | 2 (9.5) | 0 (0) |
| Platelet Count; Mean (SD); x109/L |  | 314.5 ± 440.7 | 197.1 ± 89.1 | 191.45 ± 60.9 | 254.2 ± 170.1 |
| Platelet Count; Median [IQR]; x109/L |  | 229 [161 – 333.7] | 178.5 [142.5 – 254] | 183 [141 – 246] | 237 [139 – 337] |
|  | <350 x109/L; N (%) | 228 (78.1) | 50 (94.3) | 20 (100) | 8 (88.9) |
|  | 350-500 x109/L; N (%) | 64 (21.9) | 3 (5.7) | 0 (0) | 1 (11.1) |
| Alanine Transaminase (ALT); Mean (SD); U/L |  | 63.3 ± 86.3 | 50.4 ± 66.0 | 67.2 ± 78.1 | 120.9 ± 231.6 |
| Alanine Transaminase (ALT); Median [IQR]; U/L |  | 29.8 [10.1 – 90.2] | 22.9 [8.0– 67.7] | 53 [11.6 – 75.2] | 39 [29 – 107] |
| Aspartate Transaminase (AST); Mean (SD); U/L |  | 32.1 ± 26.8 | 35.7 ± 24.5 | 43.4 ± 27.9 | 121.5 ± 265.1 |
| Aspartate Transaminase (AST); Median [IQR]; U/L |  | 23.9 [14-40.5] | 28.9 [20 – 43] | 35 [21 – 69.8] | 44 [30.5 – 63] |
| Blood Urea Nitrogen (BUN); Mean (SD); mg/dl |  | 53.1± 104.2 | 99.9 ± 290.8 | 83.1 ± 160.6 | 31.9 ± 16.3 |
| Blood Urea Nitrogen (BUN) mg/dl; Median [IQR] |  | 32 [21 – 50] | 33.5 [21.5 – 51] | 36 [26 – 60] | 30 [23 – 37] |
| Creatinine; Mean (SD); mg/dl |  | 1.5 ± 1.7 | 2.2 ± 2.6 | 2.57 ± 3.12 | 1.46 ± 1.21 |
| Creatinine; Median [IQR]; mg/dl |  | 1.1 [1.1 – 1.5] | 1.1 [0.9 – 1.8] | 1 [0.9 – 2.8] | 1.1 [0.9 – 1.6] |
| C-Reactive Protein (CRP); Mean (SD); mg/L |  | 61 ± 131 | 79 ± 192 | 85 ± 155 | 54 ± 37 |
| C-Reactive Protein (CRP); Median [IQR]; mg/L |  | 36 [24 – 57] | 37 [24 – 54] | 41 [29.5 – 62.5] | 30 [23.5 – 37] |
| Lactate Dehydrogenase (LDH); Mean (SD); U/L |  | 604.8 ± 1536.7 | 450.8 ± 443.3 | 443.0 ± 268.5 | 766 ± 812.1 |
| Lactate Dehydrogenase (LDH); Median [IQR]; U/L |  | 377 [245 - 524] | 366.5 [254.5 - 571] | 252 [230 - 664] | 453 [250 -1115] |
| Fibrinogen; Mean (SD); mg/dl |  | 463.6 ± 989.6 | 706.1 ± 418.1 | - | - |
| Fibrinogen; Median [IQR]; mg/dl |  | 223 [3.9 – 490.5] | 693 [303 - 1246] | - | - |
| D- Dimer; Mean (SD); ng/ml |  | 2654.8 ± 6429.4 | 8668.4 ± 16318.1 | - | 1080.8 ± 1376.5 |
| D- Dimer; Median [IQR]; ng/ml |  | 1027 [551 - 2200] | 2303 [584.5 - 6875] | - | - |

\* Data on patients’ disposition were sparse.

AIS was diagnosed in 323(74.8%) patients (Table 2). AIS patients were grouped based on gender and age – ≤55 versus >55 years-old, ≤65 versus >65 years-old (Supplemental Table 2, 3 and 4). Out of 323 patients with AIS, 36.2% were younger than 55 years. Patients above 55 years had higher proportion of comorbidities—hypertension, 69.3% versus 39.4%, p<0.001; ischemic heart disease, 27.3% versus 13.8%, p=0.03; atrial fibrillation, 16.1% versus 6.1%, p=0.04; and carotid stenosis, 15.2% versus 4.6%, p=0.03. In addition, older stroke patients had higher cardioembolic origin according to the TOAST criteria (31.2% versus 15.6%, p=0.05). There were differences among men and women in smoking status (9.4% versus 21.2% in men, p=0.01), chronic kidney disease (7.9% versus 16.6% in men, p=0.01), NIHSS (12.0 [5.0 – 19.0] versus 8.0 [4.0 – 16.0] in men, p<0.001), and TOAST classification (p=0.05).

**Table 2.** Neurological findings among patients with acute ischemic stroke.

| Parameters | N (%) | Parameters | N (%) |
| --- | --- | --- | --- |
| Imaging Pattern of Ischemia |  | Vascular Territory based on Imaging* |  |
| Embollic/Large Vessel Athero-thromboembolism | 206 (80.5) | Anterior Cerebral Artery Unilateral | 7 (2.2) |
| Lacunar | 26 (10.2) | Anterior Cerebral Artery Bilateral | 1 (0.3) |
| Borderzone | 23 (9.0) | Middle Cerebral Artery Unilateral | 181 (56) |
| Vasculitis | 1 (0.4) | Middle Cerebral Artery Bilateral | 5 (1.5) |
| TOAST Criteria |  | Posterior Cerebral Artery Unilateral | 33 (10.2) |
| Large Artery Atherosclerosis | 56 (32.9) | Posterior Cerebral Artery Bilateral | 4 (1.2) |
| Cardioembolism | 46 (27.1) | Posterior Inferior Cerebellar Artery | 9 (2.8) |
| Small Artery Occlusion | 17 (10.0) | Missing | 64 (19.8) |
| Other Determined Etiology | 13 (7.6) | Symptoms at Onset* |  |
| Undetermined Etiology | 38 (22.4) | Hemineglect | 1 (0.4) |
| National Institute of Health Stroke Scale (NIHSS) |  | Limb Paresis Unilateral | 168 (72.4) |
| No Stroke Symptoms (NIHSS=0) | 20 (7.5) | Limb Paresis Bilateral | 2 (0.9) |
| Minor Stroke (NIHSS of 1 to 4) | 49 (18.4) | Sensory Loss Unilateral | 26 (11.2) |
| Moderate Stroke (NIHSS of 5 to 15) | 117 (43.8) | Sensory Loss Bilateral | 1 (0.4) |
| Moderate to Severe Stroke (NIHSS of 16 to 20) | 35 (13.1) | Visual Field Loss Unilateral | 14 (6.0) |
| Severe Stroke (NIHSS 21-42) | 46 (17.2) | Visual Field Loss Bilateral | 2 (0.9) |
| Large vessel Occlusion | 126 (44.5) | Gaze Preference | 13 (5.6) |
| Mechanical Thrombectomy | 24 (7.4) | Facial Paresis | 46 (19.8) |
| Intravenous Thrombolysis | 44 (13.6) | Aphasia | 56 (24.1) |
|  |  | Dysarthria | 41 (17.7) |
|  |  | Altered Level of Consciousness | 57 (24.6) |
|  |  | Ataxia | 21 (9.1) |
|  |  | Seizure | 7 (3.0) |
|  |  | Missing | 91 (28.2) |

\*Due to multiple infarcts in some patients, the summation may exceed 100%.

The distribution of AIS subtypes according to TOAST classification was the following: large-artery atherosclerosis (33%), cardioembolism (27%), small vessel occlusion (10%), ischemic stroke of other determined etiology (8%) and ischemic stroke of undetermined etiology (22%). The subgroups of the patients according to the TOAST classification were different in terms of age, gender, the prevalence of LVO, need for mechanical ventilation, hypertension, ischemic heart disease, atrial fibrillation, carotid stenosis, chronic kidney disease, and neoplasm, the median of laboratory results (lymphocytes counts, creatinine), and imaging patterns. We observed lower median of D-dimer in patients with large artery atherosclerosis compared to those with cardio-embolism strokes (486.5 [371.5 – 1422.5] ng/ml versus 1100.0 [955.0 – 2355.0] ng/ml, p=0.04). There were no differences in terms of LDH or fibrinogen among TOAST subgroups (Table 2, Supplemental Table 5). Patients grouped by NIHSS strata were different in terms of the prevalence of LVO, intravenous thrombolysis and mechanical thrombectomy, imaging patterns, TOAST classifications, need for mechanical ventilation, ischemic heart disease, atrial fibrillation, and mean of laboratory results (white blood cell counts, alanine transaminase, aspartate transaminase, creatinine, fibrinogen, and D-dimer) (Table 3, Supplemental Table 6). Patients with LVO were different from those without LVO in terms of NIHSS, imaging patterns, TOAST criteria, and prevalence of intravenous thrombolysis, mechanical thrombectomy, and ischemic heart disease. (Table 3, Supplemental Table 7). Supplemental Tables 8 and 9 also present the difference among subgroups in terms of neuroimaging findings and the interval between SARS-CoV-2 infection and stroke. AIS patients were additionally compared based on geographical regions (Middle East, Asia, Europe, and America, Supplemental Table 10), and countries’ health expenditures (low, or middle to high, Supplemental Table 11). We observed differences among the patients in terms of comorbidities: ischemic heart disease, chronic kidney disease, and congestive heart failure with ejection fraction <40%. We also observed differences among patients when considering the median of laboratory results (white blood cell count, platelet count, lactate dehydrogenase, fibrinogen, and D-dimer), the proportion of patients who underwent mechanical thrombectomy, and neuro-imaging findings. We observed a significant difference regarding the TOAST criteria in both regional and health-expenditure subgroups; small artery occlusion in the Middle East: 18.3%, America: 5.3%, and Europe: 4.1%, p<0.001; 4% versus 18.3% in countries with lower health-expenditure, p<0.001. We further detected higher NIHSS in countries with lower health-expenditure—11.0 [5.0–17.0] versus 8.0 [3.0–17.0], p=0.02. There was no significant difference among regions and health-expenditure subgroups in terms of the prevalence of LVO.

**Table 3.** Baseline characteristics and neuroimaging findings under each outcome measures.

| Parameters | TOAST Criteria |  |  |  |  |  | Large Vessel Occlusion |  |  | National Institutes of Health Stroke Scale (NIHSS) |  |  |  |  |  |
| --- | --- | --- | --- | --- | --- | --- | --- | --- | --- | --- | --- | --- | --- | --- | --- |
|  | A: Large Artery Atherosclerosis<br>N = 56 (32.9%) | B: Cardio-embolism<br>N = 46 (27.1%) | C: Small Artery Occlusion<br>N = 17 (10.0%) | D: Other Determined<br>N = 13 (7.6%) | E: Undetermined<br>N = 38 (22.4%) | P value | A: Large Vessel Occlusion<br>N = 126 (44.5%) | B: Other Strokes<br>N = 157 (55.5%) | P value | A: No Stroke Symptoms (NIHSS = 0)<br>N = 20 (7.5%) | B: Minor Stroke (NIHSS = 1 to 4)<br>N = 49 (18.4%) | C: Moderate Stroke (NIHSS 5 to 15)<br>N = 117 (43.8%) | D: Moderate to Severe Stroke<br>(NIHSS = 16 to 20) N = 35 (13.1%) | E: Severe Stroke (NIHSS = 21 to 42)<br>N = 46 (17.2%) | P-value |
| Age; Mean (SD); Years | 63 ± 15 | 72 ± 14<br>D (0.021) | 67 ± 18 | 57 ± 16 | 65 ± 18 | 0.01 | 65.7 ± 14. | 68 ± 16. | 0.22 | 67 ± 13 | 65 ± 17 | 66 ± 15 | 66 ± 16 | 69 ± 15 | 0.77 |
| Sex; Female; N (%) | 19 (33.9) | 21 (45.7) | 3 (17.6) | 9 (69.2)<br>C (0.043) | 15 (39.5) | 0.05 | 46 (36.5) | 71 (45.2) | 0.14 | 6 (30.0) | 20 (40.8) | 44 (37.6) | 16 (45.7) | 22 (47.8) | 0.60 |
| Large Vessel Occlusion; N (%) | 46 (85.2)<br>B (0.003)<br>D (<0.001)<br>E (<0.001) | 24 (53.3) | 0 (0.0) | 2 (18.2) | 7 (20.0) | <0.001 | - | - | - | 1 (8.3) | 8 (17.0) | 49 (44.5)<br>B (0.01) | 23 (67.6)<br>A (0.004)<br>B (<0.001) | 31 (75.6)<br>A (<0.001)<br>B (<0.001)<br>C (0.01) | <0.001 |
| Intravenous Thrombolysis; N (%) | 14 (25.0) | 8 (17.4) | 0 (0.0) | 2 (15.4) | 4 (10.5) | 0.12 | 28 (22.2)<br>B (0.002) | 14 (8.9) | 0.002 | 0 (0.0) | 2 (4.1) | 25 (21.4)<br>B (0.04) | 11 (31.4)<br>B (0.004) | 5 (10.9) | <0.001 |
| Mechanical Thrombectomy; N (%) | 10 (17.9) | 10 (21.7) | 0 (0.0) | 1 (7.7) | 2 (5.3) | 0.07 | 24 (19.0) | 0 (0.0) | <0.001 | 0 (0.0) | 1 (2.0) | 8 (6.8) | 9 (25.7)<br>B (0.01)<br>C (0.01) | 6 (13.0) | <0.001 |
| National Institutes of Health Stroke Scale (NIHSS); Median [IQR] | 9.0<br>[5.0 – 17.0] | 13.0<br>[8.0 -20.0] | 4.0<br>[2.0 -8.0] | 14.0<br>[6.0 – 18.0] | 7.0<br>[3.0 – 17.0] | 0.10 | 15.0<br>[8.0 – 21.0] | 6.0<br>[3.0 – 12.0] | <0.001 | - | - | - | - | - | - |
| Imaging Patterns |  |  |  |  |  |  |  |  |  |  |  |  |  |  |  |
| Embolic/Large Vessel athero-Thromboembolism; N (%) | 54 (96.4)<br>C (<0.001)<br>D (0.017) | 43 (93.5)<br>C (<0.001) | 4 (23.5) | 9 (69.2) | 34 (89.5)<br>C (<0.001) | <0.001 | 99 (88.4)<br>B (0.003) | 99 (73.3) | <0.001 | 4 (57.1) | 31 (73.8) | 85 (81.7) | 24 (75.0) | 39 (92.9) | <0.001 |
| Lacune; N (%) | 0 (0.0) | 1 (2.2) | 13 (76.5)<br>B (<0.001)<br>D (0.001)<br>E (<0.001) | 1 (7.7) | 4 (10.5) |  | 2 (1.8) | 12 (17.0)<br>B (<0.001) |  | 2 (28.6) | 8 (19.0) | 10 (9.6) | 1 (3.1) | 1 (2.4) |  |
| Borderzone; N (%) | 2 (3.6) | 1 (2.2) | 0 (0.0) | 3 (23.1)<br>A (0.044)<br>B (0.024) | 0 (0.0) |  | 11 (9.8) | 12 (8.9) |  | 0 (0.0) | 3 (7.1) | 9 (8.7) | 7 (21.9) | 2 (4.8) |  |
| Vasculitis Pattern; N (%) | 0 (0.0) | 1 (2.2) | 0 (0.0) | 0 (0.0) | 0 (0.0) |  | 0 (0.0) | 1 (0.7) |  | 1 (14.3) | 0 (0.0) | 0 (0.0) | 0 (0.0) | 0 (0.0) |  |
| Interval Between infection Onset to Index Event; Median [IQR]; Days | 4.0<br>[1.0 – 10.0] | 2.0<br>[0.0 – 11.0] | 2.0<br>[0.0 – 6.0] | 10.0<br>[4.0 – 17.0] | 4.0<br>[0.0 – 12.0] | 0.12 | 46 (58.2)<br>A (<0.001) | 8 (9.6) | <0.001 | 3.0<br>[0.0 – 12.0] | 3.0<br>[0.0 – 10.0] | 4.0<br>[1.0 – 8.0] | 3.0<br>[0.0 – 10.0] | 1.0<br>[0.0 – 10.0] | 0.64 |
| Mechanical Ventilation; N (%) | 22 (29.3) | 10 (21.7) | 4 (23.5) | 9 (69.2) | 11 (28.9) | 0.02 | 36 (28.6) | 42 (26.8) | 0.73 | 2 (10.0) | 8 (16.3) | 25 (21.4) | 12 (34.3) | 29 (63.0)<br>A (0.001)<br>B (<0.001)<br>C (<0.001) | <0.001 |
| Disposition* |  |  |  |  |  |  | 0 (0.0) | 17 (20.5) |  |  |  |  |  |  |  |
| Discharged Home; N (%) | 24 (42.9) | 16 (34.8) | 11 (64.7) | 8 (61.5) | 9 (24.3) | 0.12 | 46 (37.7) | 66 (44.6) | 0.24 | 5 (62.5) | 28 (62.2)<br>D (0.001)<br>E (0.001) | 52 (47.3)<br>D (0.24)<br>E (0.02) | 6 (17.6) | 9 (20.5) | <0.001 |
| In Hospital Mortality; N (%) | 14 (25.0) | 13 (28.3) | 3 (17.6) | 2 (15.4) | 9 (24.3) |  | 40 (32.8) | 35 (23.6) |  | 1 (12.5) | 5 (11.1) | 18 (16.4) | 19 (55.9)<br>B (<0.001)<br>C (<0.001) | 23 (52.3)<br>B (<0.001)<br>C (<0.001) |  |
| Still in Hospital/Subacute Care; N (%) | 18 (32.1) | 17 (37.0) | 3 (17.6) | 3 (23.1) | 19 (54.1) |  | 36 (29.5) | 47 (31.8) |  | 2 (25.0) | 12 (26.7) | 40 (36.4) | 9 (26.5) | 12 (27.3) |  |
| Length of Hospital Stay; Median (IQR); Days | 6.0 [4.0 – 15.0] | 7.0 [5.0 – 14.0] | 7.0 [5.0 – 16.0] | 20.0 [4.0 – 35.0] | 8.0 [6.0 – 26.0] | 0.17 | 6.0 [4.0 – 15.0] | 8.0 [5.0 – 17.0] | 0.10 | 2 (10.0) | 8 (16.3) | 25 (21.4) | 12 (34.3) | 29 (63.0)<br>A (0.001)<br>B (<0.001)<br>C (<0.001) | <0.001 |

\* Data on patients' disposition were sparse.

A total of 91(21.1%) patients presented with the ICH (Table 4). Among them, 3(3.3%) had simultaneous SAH and IPH without any evidence of aneurysm, and 4(4.4%) were presented with simultaneous intraventricular hemorrhage and IPH. Isolated SAH occurred in 23(25.3%), and isolated IPH in 61(67%) of the patients with hemorrhagic stroke. Eighteen (4.2%) patients experienced cerebral sinus or cortical venous thrombosis (Table 3); among them, 5(27.8%) had multiple vascular involvements.

**Table 4.** Localization and presenting symptoms regarding the SARS-CoV-2 infected patients with intracranial hemorrhage and cerebral sinus and venous thrombosis.

| Localization | N (%) | Symptoms | N (%) |
| --- | --- | --- | --- |
| **Intracranial Hemorrhage** |  |  |  |
| Subarachnoid Hemorrhage | 23 (25.3) | Intracranial (Minus Subarachnoid) Hemorrhage | 68 (15.7) |
| Middle Cerebral Artery Aneurysm | 2 (8.6) | Altered Level of Consciousness | 27 (39.7) |
| Posterior Cerebral Artery Aneurysm | 1 (4.3) | Limb Paresis | 25 (36.7) |
| Anterior Communicating Artery Aneurysm | 2 (8.6) | Aphasia | 3 (4.4) |
| Basilar Top Aneurysm | 2 (8.6) | Facial Paresis | 2 (2.9) |
| No Aneurysm Detected | 16 (69.5) | Sensory Loss | 3 (2.9) |
| Subarachnoid and Intraparenchymal Hemorrhages | 3 (3.3) | Dysarthria | 4 (2.9) |
| Intraventricular and Intraparenchymal Hemorrhages | 4 (4.4) | Ataxia | 5 (2.9) |
| Intraparenchymal Hemorrhage | 61 (67) | Visual Field Defect | 1 (1.5) |
| Cerebellar Hemorrhage | 3 (7.0) | Subarachnoid Hemorrhage | 23 (25.3) |
| Brain Stem Hemorrhage | 4 (9.3) | Thunderclap headache | 16 (69.5) |
| Basal Ganglia Hemorrhage | 19 (43.1) | Decreased level of consciousness | 5 (21.7) |
| Thalamic Hemorrhage | 7 (16.3) | Cerebral herniation | 1 (4.3) |
| Lobar Hemorrhage | 20 (46.5) | Seizure | 1 (4.3) |
| Missing | 17 (28.3) | Missing | 21 (30.9) |
| **Cerebral Venous and Sinus Thrombosis** |  |  |  |
| Superior Sagittal Sinus | 6 (33.3) | Headache | 9 (50) |
| Sigmoid Sinus | 6 (33.3) | Seizure | 7 (38.9) |
| Transverse Sinus | 5 (27.8) | Altered Level of Consciousness | 5 (27.8) |
| Lateral Sinus | 3 (16.7) | Increased Intracerebral Pressure | 2 (11.1) |
| Straight Sinus | 1 (5.6) | Concomitant Intracranial Hemorrhage | 2 (11.1) |
| Cortical Veins | 1 (5.6) |  |  |
| Multiple Sinus or Venous Involvement | 5 (27.8) |  |  |

#### Clustering and Subgroup Analysis

The structure of the models and proportion of the comorbidities under each model are available in Supplemental Tables 12-14 (AIS patients) and Supplemental Tables 15-17(IPH patients). Clustering the patients into 4 or 5 subgroups (ML-K_4_, ML-S_4_, ML-K_5_, and ML-S_5_; Supplemental Tables 14A, 14B, 17A, and 17B) were not informative and not included for further comparisons. Supplemental Figures 2 (AIS) and 3 (IPH) demonstrate contingency matrices. Similarity among different models was assessed to be strong (Sim>80) in all matrices except comparisons of unsupervised algorithms (ML-K_3_ and ML-S_3_) and expert opinions (EX-A_3_ and EX-S_3_) among AIS patients— moderate similarity, Sim=66%-73%. Figure 2A visualizes the comorbidities among AIS patients. The hierarchical cluster demonstrates 4 subgroups, with a relatively large group with no or limited comorbidities. Figure 2B demonstrates comorbidities among IPH patients. Hypertension and diabetes were the most repeated and overlapped comorbidities among both AIS and IPH cohorts.

**Figure 2.**
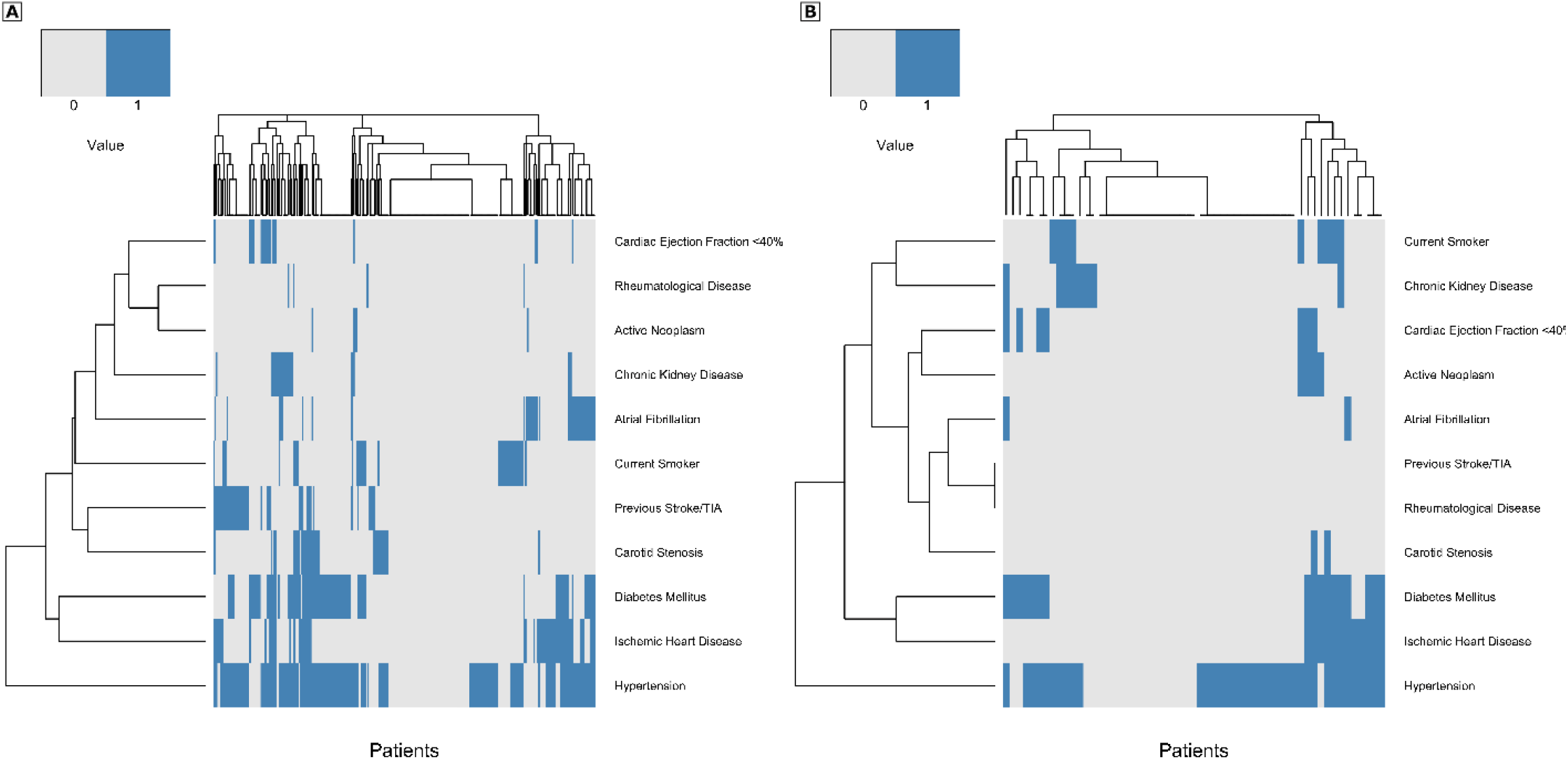
The image demonstrates the hierarchical clustering of the patients based on the presence (blue line) or absence of the comorbidities. A. SARS-CoV-2 infected patients with acute ischemic stroke. B. SARS-CoV-2 infected patients with intraparenchymal hemorrhage.

When the AIS patients were divided into two subgroups based on the existing comorbidities, all models (EX-A_2_, EX-S_2_, ML-K_2_, and ML-S_2_) presented differences among the patients according to the TOAST criteria and median of creatinine. EX-A_2_, EX-S_2_, and ML-K_2_ also showed differences in the mean of age among the subgroups (Supplemental Table 18). When patients were divided into 3 subgroups, all models demonstrate a difference in mean of age among the patients. Additionally, subclasses were different in terms of TOAST criteria (EX-A_3_, EX-S_3_, ML-S_3_), intravenous thrombolysis (ML-S_3_ and EX-A_3_), and LVO (ML-S_3_) (Supplemental Table 19).

Dividing the IPH patients into two subgroups presented differences in white blood cells’ count (ML-K_2_ and EX-A_2_) and neutrophils’ count (ML-K_2_, EX-A_2_, and EX-S_2_). We observed differences among subgroups of IPH patients in terms of age (ML-K_3_, ML-S_3_, EX-A_3_, and EX-S_3_), and ICH score (EX-A_3_ and EX-S_3_). Patients with no known comorbidities had younger age and higher ICH score in comparison with those having 1 or 2 comorbidities, or >2 comorbidities: EX-A3 model: mean age of 55±19 years compared to 69±15 years and 64±17 years, p=0.03; ICH score of 4[2-5] compared to 3[2-3] and 3[2-4], p=0.02. EX-S3 model: mean age of 53±19 years compared to 70±13 years and 63±17 years, p=0.001; ICH score of 4 [2-4] compared to 3 [2-3] and 3[2-5], p=0.02 (Supplemental Tables 20 and 21).

## Discussion

To our knowledge, this is to date the largest study that comprehensively presents the characteristics and stroke subtypes of stroke in SARS-CoV-2 infected patients at a multinational level. The results of our work indicated a relatively high number of young AIS patients, male predominance, asymptomatic SARS-CoV-2 infection in more than one-third of the AIS patients, absence of traditional stroke risk factors in about one-fourth of the AIS patients, and a low rate of small artery occlusion and lacunar infarcts. We also noted significant differences regarding the TOAST criteria in both regional and health-expenditure subgroups as well as higher NIHSS among countries with lower health-expenditure. About one third of IPH patients did not have vascular risk factors but presented with higher ICH scores.

Although the definition of young stroke is debatable, the majority of the studies considered 50 or 55 years as the cut-off.[40] We considered both 55 and 65 years-old as the cut-off and realized that younger patients had fewer comorbidity rates (Supplemental Tables 2 and 3). Out study results showed that 36% of our AIS patients were younger than 55 years old. This proportion was considerably higher than the reported proportions (12.9% to 20.7%) of young stroke patients.[48,49] In our study, 64.4% of the patients under 55 years had AIS. The median age of AIS patients in our study was 68[58-78] years, with no substantial differences across geographical regions (Supplemental Table 10). A case series from New York on 32 AIS patients with SARS-CoV-2 showed a median of 63 years for these patients which was significantly lower than AIS patients without SARS-CoV-2 in the same study and interval (median: 70 years), or historical cohort of the AIS patients presented to the center in 2019 (median: 68.5 years).[50] A multinational study on 174 AIS patients with SARS-CoV-2 infection reported a median age of 71 years. [24] A national surveillance study from the United Kingdom presented 77 SARS-CoV-2 infected patients with cerebrovascular events and reported a median age of 74 years.[14] The authors stated that the overall median age of 71 in 125 SARS-CoV-2 infected patients with cerebrovascular disorders, altered mental status, and peripheral neuropathy in this study is comparable to national data from the UK Government public health bodies over the same period. However, caution should be made when interpreting the summary results from patients with different cerebrovascular disorders.

In our clustering, all models showed differences among the subgroups in terms of the TOAST criteria. In addition, we observed the absence of traditional stroke risk factors in about one-fourth of the AIS patients (subgroup a of EX-A_3_). Comparing these patients with our stroke registry of 8,929 AIS patients (GNSIS, Geisinger NeuroScience Ischemic Stroke database) indicated a higher prevalence of patients with no known risk factor (22.0% in this study versus 11.5% in GNSIS, p<0.001). We also observed that the patients in this study were younger (60.0±18.0 years versus 62.9±17.6 years, p=0.004) and had more severe strokes (NIHSS of 8[4-22] versus 4[2-7], p<0.001). Further investigation is needed to present the etiology and possible risk factors among these patients as well as to define the underlying predispositions and pathogenic pathways occurring in SARS-CoV-2 infected individuals.

Among the AIS patients, 13.6% of patients received intravenous thrombolysis, while 7.4% underwent mechanical thrombectomy. These rates are similar to the multinational study on 174 AIS SARS-CoV-2 infected patients (12.7% thrombolysis, 6.9% thrombolysis, and thrombectomy, and 5.2% mechanical thrombectomy). [24] Although the rate of LVOs and IVT were almost the same in various regions, we observed a significant difference among the regions in the rate of mechanical thrombectomy—12.4% versus 2% in countries with lower health expenditure; Middle East: 2.6%, Asia: zero, America: 4.5%, and Europe: 21.1%. Overall, 44.5% of ischemic strokes were due to LVOs, without any age or sex predominance. This rate is comparable to a similar report from New York.[50] LVOs accounted for 24% to 46% of acute ischemic strokes.[51] When the definition of the LVOs is limited to ICA, MCA (M1 and M2), ACA (A1), PCA (P1), VA, and BA, similar to our study, the prevalence drops to 24-38%,[52,53] which was lower than our findings. We could not identify any significant difference in the risk of LVOs among various regions in our study (Supplemental Tables 10).

Stroke subtyping based on the TOAST criteria revealed a 33% large artery atherosclerosis (LAA) in our study, which is higher than reports from worldwide population-based studies (19-23%).[54,55] In the Middle East, our study presented 53.5% LAA, compared with the previously reported rate of 8-31%.[56-59] Small-vessel occlusion (SVO) accounted for stroke etiology in 10% of our patients, and analysis of neuro-imaging patterns showed 10.2% lacunar infarcts (lacune). These rates are lower than worldwide population-based studies—21-44% SVO,[55,60] and 21-30% lacune [55,61,62] (Supplemental Table 22). Likewise, we observed lower regional rates of SVO and lacune: in Europe, 4.1% SVO and 9.3% lacune, versus previous reports of 12-31% SVO [63-69]and 14-31% lacune;[65,70-72] in the North and South America, 5.3% SVO and 9.1% lacune, versus reported SVO rates of 15-18%[55,73] and 13-18% lacune;[55,62] and in the Middle Eastern countries, 18.3% SVO and 11.3% lacune, versus 20-25% SVO[56-60,74] and 19-26% lacune.[59,75,76] We could not identify any difference in imaging findings or subclasses of the TOAST criteria among SARS-CoV-2 infected patients who presented with chief complaints of stroke-related symptoms compared to patients with a prior diagnosis of SARS-CoV-2 infection. Similar to our findings, other reports on SARS-CoV-2 infected stroke patients suggested the lower rate of SVO and lacune, and a predilection of SARS-CoV-2 for inducing LVO.[77-79] Lacunar infarctions and SVO are more likely to produce milder deficits.[61,80-82] During the COVID-19 pandemic, patients with mild to moderate stroke symptoms were less likely to present at medical centers.[5,78,79,83] In addition, less severe stroke symptoms, mostly in critically ill patients or overwhelmed health centers, were more likely to be under-diagnosed. We observed a lower median NIHSS score in countries with middle to high-health expenditure—8[3-17] versus 11 [5-17] in countries with lower health expenditure. The regional difference in the severity of stroke in our study—NIHSS of 12[6-17] in the Middle East, versus 7[0-16] in America and 8[4-18] in Europe— may either indicate that the patients with less severe strokes did not present to the hospital or reflect the under-diagnosis in some countries. Future studies such as CASCADE (Call to Action: SARS-CoV-2 and Cerebrovascular DisordErs) are required to shed light on changes in stroke care protocols and hospitalization rate during the pandemic. [84]

Our study reported 91 patients with intracranial hemorrhage. Among the patients with SAH—23, no aneurysm was detected in 69.5% patients, which is higher than reported 15%(5%-34%) spontaneous SAH among patients without SARS-CoV-2 infection.[85] We observed that 27.9% of IPH patients had no vascular risk factor or comorbidities (subgroup ‘a’ of the EX-A3 model). These patients had higher ICH score and younger age in comparison with other patients with IPH.

In this study we observed 18 stroke patients with CVST; The average age of patients was 49 years, 78% were younger than 55 years old, and more than 60% were women. CVST was reported as a consequence of various viral infections such as the Varicella Zoster virus,[86-89] Influenza virus,[90] Herpes Simplex virus,[91] SARS,[92] and SARS-CoV-2.[11,93] Classically CVST is considered to occur in young adults, with the predilection of women.[94] However, the sex ratio varies widely - 44.7% to 83.4% in women.[95] A systematic review on the sex ratio of 23,638 patients with CVST demonstrated an increasing trend of women proportion (54.8% before 1981 to 69.8% after 2001), likely due to increased use of oral contraceptives.[96] Even though CVST patients in our study were younger than patients with other stroke subtypes, they were older than previously reported CVST patients without SARS-CoV-2 infection.[95-100] In addition, only 27.8% of the patients in our study had multiple sinus or venous involvement, which is considerably lower than previous reports in non-SARS-CoV-2 infected patients.[95,99,101,102] One reason might be the severe condition of the patients with multiple CVSTs that prevent the proper diagnosis of these patients.

To our knowledge, this is the largest study on stroke in patients infected with SARS-CoV-2. This work has several limitations. Despite that we included centers from multiple countries and presented a comprehensive panel of patients’ characteristics, some of the specific laboratory parameters related to rare stroke causes (e.g., antiphospholipid antibodies) were not included in this study. The collaborators tried to identify SARS-CoV-2 patients who presented with stroke as the first and only symptom, but difficulty in measuring all symptoms related to COVID-19 (such as fatigue, anosmia, and ageusia) should be taken into consideration. Due to the small sample and heterogeneity of the patients with subclasses of SAH or CVST, we did not apply the machine learning on this subgroup for further exploration. Although attempts were made to minimize the selection bias by including patients from different ethnicities, ecological conditions, and health care systems, this study may suffer from selection bias and low power in some subgroups. Further studies that include a control population are warranted.

In conclusion, we observed a relatively high number of young, and asymptomatic SARS-CoV-2 infections among stroke patients. Traditional vascular risk factors were absent among a relatively large cohort of patients. Among hospitalized patients, the stroke severity was lower and rate of mechanical thrombectomy was higher among countries with middle to high-health expenditure

## Data Availability

Available upon request

## Acknowledgement

We appreciate the efforts of all other health care providers and administrators who contributed data for this study.

## References

[1] Mao L, Jin H, Wang M, Hu Y, Chen S, He Q, et al. Neurologic Manifestations of Hospitalized Patients With Coronavirus Disease 2019 in Wuhan, China. JAMA Neurol 2020. doi:10.1001/jamaneurol.2020.1127.

[2] Li Y, Wang M, Zhou Y, Chang J, Xian Y, Mao L, et al. Acute Cerebrovascular Disease Following COVID-19: A Single Center, Retrospective, Observational Study. SSRN Electron J 2020;19. doi:10.2139/ssrn.3550025.

[3] Oxley TJ, Mocco J, Majidi S, Kellner CP, Shoirah H, Singh IP, et al. Large-Vessel Stroke as a Presenting Feature of Covid-19 in the Young We. N Engl J Med 2020. doi:10.1056/NEJMc2001272.

[4] Sweid A, Hammoud B, Weinberg JH, Oneissi M, Raz E, Shapiro M, et al. Letter: Thrombotic Neurovascular Disease in COVID-19 Patients. Neurosurgery 2020;0. doi:10.1093/neuros/nyaa254.

[5] Yaghi S, Ishida K, Torres J, Mac Grory B, Raz E, Humbert K, et al. SARS2-CoV-2 and Stroke in a New York Healthcare System. Stroke 2020:STROKEAHA120030335. doi:10.1161/STROKEAHA.120.030335.

[6] Mehrpour M, Shuaib A, Farahani M, Hatamabadi HR, Fatehi Z, Ghaffari M, et al. EXPRESS: COVID-19 and stroke in Iran; a case series, and effects on stroke admissions. Int J Stroke 2020:174749302093739. doi: 10.1177/1747493020937397.

[7] Morassi M, Bagatto D, Cobelli M, D’Agostini S, Gigli GL, Bnà C, et al. Stroke in patients with SARS-CoV-2 infection: case series. J Neurol 2020:1–8. doi:10.1007/s00415-020-09885-2.

[8] D’Anna L, Kwan J, Brown Z, Halse O, Jamil S, Kalladka D, et al. Characteristics and clinical course of Covid-19 patients admitted with acute stroke. J Neurol 2020. doi:10.1007/s00415-020-10012-4.

[9] Craen A, Logan G, Ganti L. Novel Coronavirus Disease 2019 and Subarachnoid Hemorrhage: A Case Report. Cureus 2020;12. doi:10.7759/cureus.7846.

[10] Reddy ST, Garg T, Shah C, Nascimento FA, Imran R, Kan P, et al. Cerebrovascular Disease in Patients with COVID-19: A Review of the Literature and Case Series. Case Rep Neurol 2020;77030:199–209. doi: 10.1159/000508958.

[11] Poillon G, Obadia M, Perrin M, Savatovsky J, Lecler A. Cerebral venous thrombosis associated with COVID-19 infection: Causality or coincidence? J Neuroradiol 2020. doi:10.1016/j.neurad.2020.05.003.

[12] Zhang Y, Xiao M, Zhang S, Xia P, Cao W, Jiang W, et al. Coagulopathy and Antiphospholipid Antibodies in Patients with Covid-19. N Engl J Med 2020;382:e38. doi: 10.1056/nejmc2007575.

[13] Escalard S, Maïer B, Redjem H, Delvoye F, Hébert S, Smajda S, et al. Treatment of Acute Ischemic Stroke due to Large Vessel Occlusion With COVID-19. Stroke 2020:1–4. doi:10.1161/strokeaha.120.030574.

[14] Varatharaj A, Thomas N, Ellul MA, Davies NWS, Pollak TA, Tenorio EL, et al. Neurological and neuropsychiatric complications of COVID-19 in 153 patients: a UK-wide surveillance study. The Lancet Psychiatry 2020;2:1–8. doi:10.1016/S2215-0366(20)30287-X.

[15] Barrios-López JM, Rego-García I, Muñoz Martínez C, Romero-Fabrega JC, Rivero Rodríguez M, Ruiz Giménez JA, et al. Ischaemic stroke and SARS-CoV-2 infection: a causal or incidental association? Neurol (English Ed 2020. doi:10.1016/j.nrleng.2020.05.008.

[16] Klok FA, Kruip MJHA, van der Meer NJM, Arbous MS, Gommers DAMPJ, Kant KM, et al. Incidence of thrombotic complications in critically ill ICU patients with COVID-19. Thromb Res 2020. doi:10.1016/j.thromres.2020.04.013.

[17] García Espinosa J, Moya Sánchez E, Martínez Martínez A. Severe COVID-19 with debut as bilateral pneumonia, ischemic stroke, and acute myocardial infarction. Med Clin (Barc) 2020. doi:10.1016/j.medcli.2020.04.012.

[18] Gomez-Arbelaez D, Ibarra-Sanchez G, Garcia-Gutierrez A, Comanges-Yeboles A, Ansuategui-Vicente M, Gonzalez-Fajardo JA. Covid-19-Related Aortic Thrombosis: a Report of Four Cases. Ann Vasc Surg 2020. doi:10.1016/j.avsg.2020.05.031.

[19] Viguier A, Delamarre L, Duplantier J, Olivot JM, Bonneville F. Acute ischemic stroke complicating common carotid artery thrombosis during a severe COVID-19 infection. J Neuroradiol 2020. doi:10.1016/j.neurad.2020.04.003.

[20] Malentacchi M, Gned D, Angelino V, Demichelis S, Perboni A, Veltri A, et al. Concomitant brain arterial and venous thrombosis in a COVID-19 patient. Eur J Neurol 2020:0–3. doi:10.1111/ene.14380.

[21] Valderrama EV, Humbert K, Lord A, Frontera J, Yaghi S. Severe Acute Respiratory Syndrome Coronavirus 2 Infection and Ischemic Stroke. Stroke 2020:STROKEAHA120030153. doi:10.1161/STROKEAHA.120.030153.

[22] Tunç A, Ünlübaş Y, Alemdar M, Akyüz E. Coexistence of COVID-19 and acute ischemic stroke report of four cases. J Clin Neurosci Off J Neurosurg Soc Australas 2020. doi:10.1016/j.jocn.2020.05.018.

[23] Avula A, Nalleballe K, Narula N, Sapozhnikov S, Dandu V, Toom S, et al. COVID-19 presenting as stroke. Brain Behav Immun 2020. doi: 10.1016/j.bbi.2020.04.077.

[24] Ntaios G, Michel P, Georgiopoulos G, Guo Y, Li W, Xiong J, et al. Characteristics and Outcomes in Patients With COVID-19 and Acute Ischemic Stroke The Global COVID-19 Stroke Registry. Stroke 2020. doi:10.1016/j.amjcard.2019.02.021.

[25] Shahjouei S, Naderi S, Li J, Khan A, Chaudhary D, Farahmand G, et al. Risk of Cerebrovascular Events in Hospitalized Patients with SARS-CoV-2 Infection. Ebiomedicine 2020. doi:http://dx.doi.org/10.2139/ssrn.3605289.

[26] Ver Hage A. The NIH stroke scale: a window into neurological status. Com Nurs Spectr 2011;24:44–49.

[27] Adams H., Bendixen B., Kappelle L., Biller J, Love B., Gordon D., et al. Classification of Subtype of Acute Ischemic Stroke. Stroke 1993;23:35–41. doi:10.1161/01.STR.24.1.35.

[28] von Elm E, Altman DG, Egger M, Pocock SJ, Gøtzsche PC, Vandenbroucke JP. The strengthening the reporting of observational studies in epidemiology (STROBE) statement: Guidelines for reporting observational studies. Int J Surg 2014;12:1495–1499. doi:10.1016/j.ijsu.2014.07.013.

[29] Equator Network. Enhancing the QUAlity and Transparency Of health Research. Univ Oxford 2019. https://www.equator-network.org.

[30] Sacco RL, Kasner SE, Broderick JP, Caplan LR, Connors JJB, Culebras A, et al. An updated definition of stroke for the 21st century: a statement for healthcare professionals from the American Heart Association/American Stroke Association. Stroke 2013;44:2064–2089. doi:https://dx.doi.org/10.1161/STR.0b013e318296aeca.

[31] WHO. Laboratory testing for coronavirus disease 2019 (COVID-19) in suspected human cases. Interim Guid 2020:1–7.

[32] Prabhakaran S, Silver AJ, Warrior L, McClenathan B, Lee VH. Misdiagnosis of Transient Ischemic Attacks in the Emergency Room. Cerebrovasc Dis 2008;26:630–635. doi:10.1159/000166839.

[33] Sadighi A, Stanciu A, Banciu M, Abedi V, Andary N El, Holland N, et al. Rate and associated factors of transient ischemic attack misdiagnosis. ENeurologicalSci 2019;15:100193. doi:10.1016/j.ensci.2019.100193.

[34] Nadarajan V, Perry RJ, Johnson J, Werring DJ. Transient ischaemic attacks: mimics and chameleons. Pract Neurol 2014;14:23–31. doi:https://dx.doi.org/10.1136/practneurol-2013-000782.

[35] Potter GM, Marlborough FJ, Wardlaw JM. Wide variation in definition, detection, and description of lacunar lesions on imaging. Stroke 2011. doi:10.1161/STROKEAHA.110.594754.

[36] Wessels T, Röttger C, Jauss M, Kaps M, Traupe H, Stol E. Identification of embolic stroke patterns by diffusion-weighted MRI in clinically defined lacunar stroke syndromes. Stroke 2005. doi:10.1161/01.STR.0000158908.48022.d7.

[37] Bang OY, Ovbiagele B, Liebeskind DS, Restrepo L, Yoon SR, Saver JL. Clinical determinants of infarct pattern subtypes in large vessel atherosclerotic stroke. J Neurol 2009;256:591–599. doi: 10.1007/s00415-009-0125-x.

[38] Abdel Razek AAK, Alvarez H, Bagg S, Refaat S, Castillo M. Imaging spectrum of CNS vasculitis. Radiographics 2014;34:873–894. doi:10.1148/rg.344135028.

[39] Waqas M, Mokin M, Primiani CT, Gong AD, Rai HH, Chin F, et al. Large Vessel Occlusion in Acute Ischemic Stroke Patients: A Dual-Center Estimate Based on a Broad Definition of Occlusion Site. J Stroke Cerebrovasc Dis 2020;29:104504.

[40] F. C. Defining Young Stroke. YoungStroke, Inc 2017:1–15.

[41] Global Health Expenditure Database n.d. https://apps.who.int/nha/database/Select/Indicators/en (accessed June 30, 2020).

[42] IBM. Downloading IBM SPSS Statistics 26. Ibm 2020.

[43] Von Luxburg U. A tutorial on spectral clustering. Stat Comput 2007;17:395–416. doi:10.1007/s11222-007-9033-z.

[44] Palacio-Niño J-O, Berzal F. Evaluation Metrics for Unsupervised Learning Algorithms. ArXiv 2019:arXiv-1905.

[45] Andy Bunn MK. An Introduction to dplR. Ind Commer Train 2008;10:11–18. doi:10.1108/eb003648.

[46] Warnes GR, Bolker B, Bonebakker L, Gentleman R, Huber W, Liaw A, et al. gplots: Various R programming tools for plotting data. R Packag Version 2009;2:1.

[47] Pedregosa F, Varoquaux G, Gramfort A, Michel V, Thirion B, Grisel O, et al. Scikit-learn: Machine learning in Python. J Mach Learn Res 2011;12:2825–2830.

[48] Cabral NL, Freire AT, Conforto AB, Dos Santos N, Reis FI, Nagel V, et al. Increase of stroke incidence in young adults in a middle-income country a 10-year population-based study. Stroke 2017;48:2925–2930. doi: 10.1161/STROKEAHA.117.018531.

[49] Kissela BM, J.C. K, Alwell K, Moomaw JC, Woo D, Adeoye O, et al. Age at stroke. Neurol 1781-1787 2012;79:1781–1787.

[50] Yaghi S, Ishida K, Torres J, Grory B Mac, Raz E, Humbert K, et al. SARS2-CoV-2 and Stroke in a New York Healthcare System 2020:1–10. doi:10.1161/STROKEAHA.120.030335.

[51] Rennert RC, Wali AR, Steinberg JA, Santiago-Dieppa DR, Olson SE, Pannell JS, et al. Epidemiology, Natural History, and Clinical Presentation of Large Vessel Ischemic Stroke. Clin Neurosurg 2019;85:S4–S8. doi: 10.1093/neuros/nyz042.

[52] Malhotra K, Gornbein J, Saver JL. Ischemic strokes due to large-vessel occlusions contribute disproportionately to stroke-related dependence and death: A review. Front Neurol 2017;8:1–5. doi:10.3389/fneur.2017.00651.

[53] Dozois A, Hampton L, Kingston CW, Lambert G, Porcelli TJ, Sorenson D, et al. Plumber study (prevalence of large vessel occlusion strokes in mecklenburg county emergency response). Stroke 2017;48:3397–3399. doi: 10.1161/STROKEAHA.117.018925.

[54] Ornello R, Degan D, Tiseo C, Di Carmine C, Perciballi L, Pistoia F, et al. Distribution and temporal trends from 1993 to 2015 of ischemic stroke subtypes a systematic review and meta-analysis. Stroke 2018;49:814–819. doi: 10.1161/STROKEAHA.117.020031.

[55] O’Donnell MJ, Denis X, Liu L, Zhang H, Chin SL, Rao-Melacini P, et al. Risk factors for ischaemic and intracerebral haemorrhagic stroke in 22 countries (the INTERSTROKE study): A case-control study. Lancet 2010;376:112–123. doi:10.1016/S0140-6736(10)60834-3.

[56] Saber H, Thrift AG, Kapral MK, Shoamanesh A, Amiri A, Farzadfard MT, et al. Incidence, recurrence, and long-term survival of ischemic stroke subtypes: A population-based study in the Middle East. Int J Stroke 2017;12:835–843. doi:10.1177/1747493016684843.

[57] Khealani BA, Khan M, Tariq M, Malik A, Siddiqi AI, Awan S, et al. Ischemic strokes in Pakistan: Observations from the national acute ischemic stroke database. J Stroke Cerebrovasc Dis 2014;23:1640–1647. doi:10.1016/j.jstrokecerebrovasdis.2014.01.009.

[58] Khorvash F, Khalili M, Rezvani Habibabadi R, Sarafzadegan N, Givi M, Roohafza H, et al. Comparison of acute ischemic stroke evaluation and the etiologic subtypes between university and nonuniversity hospitals in Isfahan. Iran. Int J Stroke 2019;14:613–619. doi:10.1177/1747493019828648.

[59] Lutski M, Zucker I, Shohat T, Tanne D. Characteristics and outcomes of young patients with first-ever ischemic stroke compared to older patients: The national acute stroke Israeli registry. Front Neurol 2017;8. doi:10.3389/fneur.2017.00421.

[60] Rukn S Al, Mazya M V., Hentati F, Sassi S Ben, Nabli F, Said Z, et al. Stroke in the Middle-East and North Africa: A 2-year prospective observational study of stroke characteristics in the region—Results from the Safe Implementation of Treatments in Stroke (SITS)–Middle-East and North African (MENA). Int J Stroke 2019;14:715–722. doi: 10.1177/1747493019830331.

[61] M. P, L. N, P. D, B. L. Outcomes from ischemic stroke subtypes classified by the Oxfordshire Community Stroke Project: A systematic review. Eur J Phys Rehabil Med 2011;47:19–23.

[62] O’Donnell MJ, Chin SL, Rangarajan S, Xavier D, Liu L, Zhang H, et al. Global and regional effects of potentially modifiable risk factors associated with acute stroke in 32 countries (INTERSTROKE): a case-control study. Lancet 2016;388:761–775. doi:10.1016/S0140-6736(16)30506-2.

[63] Li L, Yiin GS, Geraghty OC, Schulz UG, Kuker W, Mehta Z, et al. Incidence, outcome, risk factors, and long-term prognosis of cryptogenic transient ischaemic attack and ischaemic stroke: A population-based study. Lancet Neurol 2015;14:903–913. doi:10.1016/S1474-4422(15)00132-5.

[64] Yesilot Barlas N, Putaala J, Waje-Andreassen U, Vassilopoulou S, Nardi K, Odier C, et al. Etiology of first-ever ischaemic stroke in European young adults: The 15 cities young stroke study. Eur J Neurol 2013;20:1431–1439. doi:10.1111/ene.12228.

[65] Ihle-Hansen H, Thommessen B, Wyller TB, Engedal K, Fure B. Risk factors for and incidence of subtypes of ischemic stroke. Funct Neurol 2012;27:35–40.

[66] Hauer AJ, Ruigrok YM, Algra A, van Dijk EJ, Koudstaal PJ, Luijckx G-J, et al. Age-Specific Vascular Risk Factor Profiles According to Stroke Subtype. J Am Heart Assoc 2017;6:e005090. doi: 10.1161/JAHA.116.005090.

[67] Carrera E, Maeder-Ingvar M, Rossetti AO, Devuyst G, Bogousslavsky J. Trends in Risk Factors, Patterns and Causes in Hospitalized Strokes over 25 Years: The Lausanne Stroke Registry. Cerebrovasc Dis 2007;24:97–103. doi: 10.1159/000103123.

[68] Grau AJ, Weimar C, Buggle F, Heinrich A, Goertler M, Neumaier S, et al. Risk factors, outcome, and treatment in subtypes of ischemic stroke: The German stroke data bank. Stroke 2001;32:2559–2566. doi:10.1161/hs1101.098524.

[69] Wafa HA, Wolfe CDA, Rudd A, Wang Y. Long-term trends in incidence and risk factors for ischaemic stroke subtypes: Prospective population study of the South London Stroke Register. PLoS Med 2018;15:1–16. doi:10.1371/journal.pmed.1002669.

[70] Czlonkowska A, Ryglewicz D, Weissbein T, Baranska-Gieruszczak M, Hier DB. A prospective community-based study of stroke in warsaw, poland. Stroke 1994;25:547–551. doi:10.1161/01.STR.25.3.547.

[71] Alzamora MT, Sorribes M, Heras A, Vila N, Vicheto M, Forés R, et al. Ischemic stroke incidence in Santa Coloma de Gramenet (ISISCOG), Spain. A community-based study. BMC Neurol 2008;8:5. doi: 10.1186/1471-2377-8-5.

[72] Di Carlo A, Lamassa M, Baldereschi M, Pracucci G, Consoli D, Wolfe CDA, et al. Risk factors and outcome of subtypes of ischemic stroke. Data from a multicenter multinational hospital-based registry. The European Community Stroke Project. J Neurol Sci 2006;244:143–150. doi:10.1016/j.jns.2006.01.016.

[73] Porcello Marrone LC, Diogo LP, De Oliveira FM, Trentin S, Scalco RS, De Almeida AG, et al. Risk factors among stroke subtypes in Brazil. J Stroke Cerebrovasc Dis 2013;22:32–35. doi:10.1016/j.jstrokecerebrovasdis.2011.05.022.

[74] Senel GB, Elmali AD, Mehrvar K, Farhoudi M, Aboutalebi M, Rezaei M, et al. A survey from Turkey and Iran on comparison of risk factors and etiology in ischemic stroke. Iran J Neurol 2020;18:176–178. doi:10.18502/ijnl.v18i4.2189.

[75] Kumral E, Özkaya B, Şagduyu A, §irin H, Vardarli E, Pehliva M. The Ege Stroke Registry: A hospital-based study in the Aegean Region, Izmir, Turkey. Cerebrovasc Dis 1998;8:278–288. doi: 10.1159/000015866.

[76] Ghandehari K, Izadi-Mood Z. Khorasan stroke registry: Analysis of 1392 stroke patients. Arch Iran Med 2007;10:327–334. doi:07103/AIM.009.

[77] Oxley TJ, Mocco J, Majidi S, Kellner CP, Shoirah H, Singh IP, et al. Large-Vessel Stroke as a Presenting Feature of Covid-19 in the Young. N Engl J Med 2020;382:e60. doi:10.1056/NEJMc2009787.

[78] Jain R. Evolving Neuroimaging Findings during COVID-19. Am J Neuroradiol 2020;41:1–2. doi: 10.3174/ajnr.a6658.

[79] Siegler JE, Heslin ME, Thau L, Smith A, Jovin TG. Falling stroke rates during COVID-19 pandemic at a comprehensive stroke center: Cover title: Falling stroke rates during COVID-19. J Stroke Cerebrovasc Dis 2020;29:104953. doi:10.1016/j.jstrokecerebrovasdis.2020.104953.

[80] Favate AS, Younger DS. Epidemiology of Ischemic Stroke. Neurol Clin 2016;34:967–980. doi:10.1016/j.ncl.2016.06.013.

[81] Wilterdink JL, Bendixen B, Adams HP, Woolson RF, Clarke WR, Hansen MD. Effect of prior aspirin use on stroke severity in the Trial of Org 10172 in Acute Stroke Treatment (TOAST). Stroke 2001;32:2836–2840. doi:10.1161/hs1201.099384.

[82] Pittock SJ, Meldrum D, Hardiman O, Thornton J, Brennan P, Moroney JT. The Oxfordshire Community Stroke Project Classification: Correlation with imaging, associated complications, and prediction of outcome in acute ischemic stroke. J Stroke Cerebrovasc Dis 2003;12:1–7. doi:10.1053/jscd.2003.7.

[83] Hoyer C, Ebert A, Huttner HB, Puetz V, Kallmünzer B, Barlinn K, et al. Acute Stroke in Times of the COVID-19 Pandemic: A Multicenter Study. Stroke 2020:STROKEAHA120030395. doi:10.1161/STROKEAHA.120.030395.

[84] Abootalebi S, Aertker BM, Andalibi MS, Asdaghi N, Aykac O, Azarpazhooh MR, et al. Call to Action: SARS-CoV-2 and CerebrovAscular DisordErs (CASCADE). J Stroke Cerebrovasc Dis 2020;29. doi:10.1016/j.jstrokecerebrovasdis.2020.104938.

[85] Kim YW, Lawson MF, Hoh BL. Nonaneurysmal subarachnoid hemorrhage: An update. Curr Atheroscler Rep 2012;14:328–334. doi:10.1007/s11883-012-0256-x.

[86] Sujay Sada, Anjaneyulu Kammineni, Meena A. Kanikannan JA. Cerebral sinus venous thrombosis: A rare complication of primary Varicella zoster virus. Neurol India 2012;60:645–646.

[87] Imam SF, Lodhi O ul haq, Fatima Z, Nasim S, Malik WT, Saleem MS. A Unique Case of Acute Cerebral Venous Sinus Thrombosis Secondary to Primary Varicella Zoster Virus Infection. Cureus 2017;9. doi:10.7759/cureus.1693.

[88] Siddiqi SA, Nishat S, Kanwar D, Ali F, Azeemuddin M, Wasay M. Cerebral venous sinus thrombosis: Association with primary varicella zoster virus infection. J Stroke Cerebrovasc Dis 2012;21:917.e1–917.e4. doi:10.1016/j.jstrokecerebrovasdis.2012.04.013.

[89] Sudhaker B, Dnyaneshwar MP, Jaidip CR, Rao SM. Cerebral venous sinus thrombosis (CVST) secondary to varicella induced hypercoagulable state in a adult. Intern Med Insid 2014;2:1. doi:10.7243/2052-6954-2-1.

[90] Taniguchi D, Nakajima S, Hayashida A, Kuroki T, Eguchi H, Machida Y, et al. Deep cerebral venous thrombosis mimicking influenza-associated acute necrotizing encephalopathy: A case report. J Med Case Rep 2017;11:10–15. doi:10.1186/s13256-017-1444-7.

[91] Leite J, Ribeiro A, Gonçalves D, Sargento-Freitas J, Trindade L, Duque V. Cerebral Venous Thrombosis as Rare Presentation of Herpes Simplex Virus Encephalitis. Case Rep Infect Dis 2019;2019:1–5. doi: 10.1155/2019/7835420.

[92] Krayenbuehl H. Thrombosis of the cerebral veins and sinus. Acta Neurochir Suppl (Wien) 1961;Suppl 7:248–254.

[93] Dahl-Cruz F, Guevara-Dalrymple N, López-Hernández N. [Cerebral venous thrombosis and SARS-CoV-2 infection]. Rev Neurol 2020;70:391–392. doi:10.33588/rn.7010.2020204.

[94] Luo Y, Tian X, Wang X. Diagnosis and Treatment of Cerebral Venous Thrombosis: A Review. Front Aging Neurosci 2018;10. doi:10.3389/fnagi.2018.00002.

[95] Gunes HN, Cokal BG, Guler SK, Yoldas TK, Malkan UY, Demircan CS, et al. Clinical associations, biological risk factors and outcomes of cerebral venous sinus thrombosis. J Int Med Res 2016;44:1454–1461. doi:10.1177/0300060516664807.

[96] Zuurbier SM, Middeldorp S, Stam J, Coutinho JM. Sex differences in cerebral venous thrombosis: A systematic analysis of a shift over time. Int J Stroke 2016;11:164–170. doi:10.1177/1747493015620708.

[97] Dentali F, Poli D, Scoditti U, di Minno MND, Stefano VD, Siragusa S, et al. Long-term outcomes of patients with cerebral vein thrombosis: A multicenter study. J Thromb Haemost 2012;10:1297–1302. doi: 10.1111/j.1538-7836.2012.04774.x.

[98] Coutinho JM, Zuurbier SM, Aramideh M, Stam J. The incidence of cerebral venous thrombosis: A cross-sectional study. Stroke 2012;43:3375–3377. doi:10.1161/STROKEAHA.112.671453.

[99] Sidhom Y, Mansour M, Messelmani M, Derbali H, Fekih-Mrissa N, Zaouali J, et al. Cerebral venous thrombosis: clinical features, risk factors, and long-term outcome in a Tunisian cohort. J Stroke Cerebrovasc Dis 2014;23:1291–1295.

[100] Ferro JM, Aguiar de Sousa D. Cerebral Venous Thrombosis: an Update. Curr Neurol Neurosci Rep 2019;19:1–9. doi:10.1007/s11910-019-0988-x.

[101] Sassi S Ben, Touati N, Baccouche H, Drissi C, Romdhane N Ben, Hentati F. Cerebral venous thrombosis: a tunisian monocenter study on 160 patients. Clin Appl Thromb 2017;23:1005–1009.

[102] Janghorbani M, Zare M, Saadatnia M, Mousavi SA, Mojarrad M, Asgari E. Cerebral vein and dural sinus thrombosis in adults in Isfahan, Iran: Frequency and seasonal variation. Acta Neurol Scand 2008; 117:117–121. doi: 10.1111/j.1600-0404.2007.00915.x.

